# Postoperative Chemoradiotherapy causes Less Fibrosis and Better Anorectal Function compared with preoperative: A retrospective study in Rectal Cancer

**DOI:** 10.1101/2024.11.28.24318150

**Authors:** Ming Zhang, Shuai Li, Guan Yu Yu, Le Qi Zhou, Hai Di Lu, Hai Feng Gong, Lian Jie Liu, Zheng Lou, Li Qiang Hao, Fu Shen, Xian Hua Gao, Wei Zhang, Yue Yu

## Abstract

**BACKGROUND:** Radiotherapy with or without chemotherapy has been widely used to improve patient outcomes due to locally advanced rectal cancer. The differential degree of pelvic fibrosis and anorectal function after preoperative or postoperative chemoradiotherapy has not been studied previously.

**METHODS:** Data of patients who received chemoradiotherapy and radical resection of rectal cancer in our hospital from January 2000 to Aug 2021 were retrospectively analyzed. Anastomotic fibrosis scores and perirectal fibrosis scores based on magnetic resonance imaging findings were used to evaluate the extent of fibrosis one year postoperatively. The overall level of anorectal function and percentage of patients eligible for ileostomy reversal were assessed two years postoperatively.

**RESULTS:** 1331 patients were finally included, with 522 and 809 patients in in the preoperative and postoperative chemoradiotherapy groups, respectively. The postoperative chemoradiotherapy group had a higher percentage of patients undergoing ileostomy reversal and had lower anastomotic fibrosis scores, perirectal fibrosis scores, Wexner scores, and percentage of patients with temporary ileostomy than the preoperative group; this difference persisted after propensity score matching. Univariate and multivariate analyses demonstrated that the timing of chemoradiotherapy was an independent predictor of the anastomotic fibrosis score, perirectal fibrosis score, percentage of patients with temporary ileostomy, and percentage of patients eligible for ileostomy reversal.

**CONCLUSIONS:** Postoperative chemoradiotherapy is associated with less pelvic fibrosis and better anorectal function than preoperative chemoradiotherapy in the treatment of patients with locally advanced rectal cancer.

## INTRODUCTION

Radiotherapy with or without chemotherapy has been widely used to improve outcomes of patients with locally advanced rectal cancer (LARC) (1). Postoperative chemoradiotherapy (CRT) has been proven to reduce the cumulative incidence of local recurrence (2). In the 1990s, the National Cancer Institute Consensus Conference concluded that the CRT should be the standard postoperative treatment for patients with pT3/T4 or N1-2 stage rectal cancer.

Patients undergoing preoperative radiotherapy have more intact blood vessels and higher oxygenation status compared to those undergoing postoperative radiotherapy, which can contribute to higher radiosensitivity, and increased rates of resectability and sphincter preservation (3). As preoperative CRT is associated with improved local control, reduced toxicity, higher tolerance, and a greater incidence of sphincter preservation than postoperative CRT (1, 4, 5), it was recommended as the preferred treatment for patients with LARC (1, 6). However, it has substantial drawbacks such as higher peri-operative mortality, higher rates of anastomotic leak and sexual problems, and more extensive negative quality of life effects (1–4). Numerous randomized controlled trials (RCTs) and meta-analyses have demonstrated that there are no significant differences in overall survival (OS) or disease-free survival (DFS) whether CRT is delivered preoperatively or postoperatively (1, 5, 7). However, the clinical complete response rate after preoperative CRT remains unsatisfactory, ranging from 5% to 30% (9).

Clinically, perirectal lymph nodes with diameter >10mm and uneven margins are considered suspicious. However, assessing lymph node metastasis solely based on lymph node size is suboptimal, with only 60%-80% accuracy, leading to unnecessary neoadjuvant radiotherapy in patients with false positive magnetic resonance imaging (MRI) lymph node results.

Radiotherapy may lead to long-term anorectal sequelae, such as fecal incontinence, urgency, clustering of stools, and increased bowel movement (8). Although the underlying mechanism of CRP-induced anorectal sequelae remain elusive, possible causes include damage to the external anal sphincter and CRT-induced fibrosis (9). CRT-induced fibrosis is a severe long-term complication caused by the persistent activation of myofibroblasts and hyperplastic remodeling of the extracellular matrix. It has been shown to lead to multiple clinical manifestations such as rectal stenosis, reduced rectal capacity, and impaired quality of life (10, 11). However, no diagnostic test is available to quantify the extent of CRT-induced fibrosis.

Preoperative CRT was assumed to preserve better anorectal function than postoperative CRT since the irradiated rectum is subsequently resected (12). However, in our experience, patients undergoing postoperative CRT had less pelvic fibrosis and more preserved anorectal function compared to those undergoing preoperative CRT. Most studies comparing preoperative and postoperative CRT have concentrated on oncological results, tolerance, acute toxicity, and sphincter preservation rate (4). No study has investigated the differences in pelvic fibrosis and anorectal function between both groups. Therefore, this study aimed to compare levels of pelvic fibrosis and anorectal function in patients with LARC undergoing preoperative or postoperative CRT.

## METHODS

### Patients

The data of patients who underwent CRT and radical resection of rectal cancer in our hospital from January 2000 to Aug 2021 were retrospectively analyzed. The study was approved by the ethics committee of the xx Hospital. The Unique Identifying number or registration ID is researchregist xxxx. Written informed consent was obtained from all the patients before commencing the study.

### Inclusion criteria were as follows

(1) Pathologically confirmed rectal adenocarcinoma;
(2) LARC diagnosis (T3-4 or N+);
(3) Pelvic MRI demonstrating the inferior margin of the rectal cancer was located within 10 cm of the anal verge;
(4) History of either preoperative or postoperative CRT;
(5) History of radical resection of rectal cancer in our hospital;
(6) History of sphincter-preserving surgery with or without temporary ileostomy;
(7) Follow up for at least two years, including viewable pelvic MRI.

### Exclusion criteria were as follows

(1) Recurrence or metastasis within the first two years after radical resection;
(2) Radical resection performed in other hospitals;
(3) Removal of rectal tumor by local excision or abdominoperineal resection;
(4) Synchronous or metachronous colorectal cancer (CRC);
(5) Any type of hereditary CRC syndrome, such as Lynch syndrome or familial adenomatous polyposis;
(6) History of both preoperative and postoperative CRT.

### Preoperative and postoperative CRT

Although all patients with T3-4 or N+ rectal cancer were recommended to undergo preoperative CRT, some declined and underwent postoperative CRT if postoperative pathology confirmed T3-4 or N+ rectal cancer. Therefore, we do not expect significant selection bias between the two groups.

Preoperative CRT was performed with three-dimensional (3D) conformal radiation therapy, with a total dose of 50.0 Gy in 2.0-Gy daily fractions. Concurrent chemotherapy consisted of capecitabine monotherapy. The interval between preoperative CRT and surgery was generally six to eight weeks. For patients who did not receive preoperative CRT, postoperative CRT was recommended if qualifying pathology was present and started approximately one to three months after radical resection. Adjuvant 5-FU-based chemotherapy was given to the postoperative CRT group. Regardless of group, CRT duration was six months.

### Analysis of MR Images

MR images were evaluated by two radiologists (S.L. and HD.L. with 8 and 10 years of experience in medical image interpretation, respectively), who were blinded to the clinical data. If the two interpretations differed, a third senior radiologist with 14 years’ experience in medical image interpretation provided the definitive interpretation. **Table S1** shows the interobserver agreement for AFS (anastomotic fibrosis score) and PFS (perirectal fibrosis score) among the three radiologists. Systematic training before assessment was referred to the MR fibrosis scoring standards. The radiological characteristics of fibrosis and adhesion in post-treatment MR images vary significantly. Typically, they consist of low-signal intensity on both T2WI and DWI, with mild-to-moderate enhancement on CE-T1WI images. Edema or effusion in the perirectal area mainly appears as high-signal intensity on both non-contrast T2-weighted and DW images.

### Definition of AFS and PFS

As described in our earlier research(13), AFS and PFS were used to grade anastomotic and perirectal fibrosis using postoperative high-resolution oblique-axial high-resolution T2WI images without fat suppression performed one year after radical resection (**Figure S1**). AFS was used to describe the extent of fibrotic proliferation, an inner thick layer or nodular focus of homogeneous intermediate-low signal intensity based on T2WI with or without stenosis, at the anastomosis site. AFS was graded as follows: 0 (thickened scar occupying < 1/3 of the anastomosis), 1 (thickened scar occupying 1/3–2/3 of the anastomosis), and 2 (thickened scar occupying more than 2/3 of the anastomosis) (**Figure 1A**). PFS was utilized to describe the extent of fibrosis and adhesion in the following parts of the pelvis: the peri-anastomotic area, presacral area, and pelvic wall. The postoperative perirectal-regional fibrotic tissue typically showed low-signal intensity on T2WI that included irregular linear or fibrous appearances, with or without adhesion of surrounding tissues. PFS was graded as follows:0 (no perirectal fibrosis and adhesion), 1 (fibrosis and adhesion were limited to one part of the pelvis), 2 (fibrosis and adhesion could be found in two or three parts of the pelvis) (**Figure 1B**).

**Figure 1.** Examples of anastomotic fibrosis score (AFS) and perirectal fibrosis score (PFS) in schematic diagram and postoperative rectal MR images (based on high-resolution oblique-axial high-resolution T2WI without fat suppression)(13). A: AFS=0, no fibrosis on the anastomosis (a low-signal intensity around the rectal wall, arrow). B: AFS=1, the anastomosis has some thickened fibrosis at the right posterior rectal wall (about 1/3 circle, arrow). C: AFS=2, the anastomosis was almost occupied by the thickened fibrosis (about 7/8 circle, arrow). D: PFS=0, no perirectal fibrosis and adhesion in the peri-anastomotic area (arrow), presacral area and pelvic wall. E: PFS=1, fibrosis at the right levator ani muscle (arrow). F: PFS=2, fibrosis and adhesion at the peri-anastomotic area, presacral area, and pelvic wall (arrow).

### Follow-up

After surgery, follow-up was performed every three months during the first two years, then every six months for another three years, and annually thereafter. Colonoscopy was also performed annually. In addition, ileostomy reversal and anorectal function (Wexner fecal incontinence score) were investigated approximately two years after surgery (**Figure S1**) (14).

### Definition of included variables

Tumor height was defined as the distance between the lower edge of the tumor and the anal verge by colonoscope. The preoperative TNM stage (cTNM) and postoperative pathological TNM stage (pTNM) were classified according to the eighth edition of the American Joint Committee on Cancer (AJCC) cancer staging system. Age was defined as “the age at the time of rectal cancer diagnosis”.

### Statistical analysis

Categorical variables were described as frequency and percentage, and compared with the chi-square test. The Fisher’s exact test was used when the expected value was less than five. Normally distributed continuous variables were expressed as mean ± standard deviation (SD), and compared with the student’s t-test.

Propensity score matching (PSM) was used to balance the factors that may affect the timing of CRT and Wexner scores, including age, gender, body mass index (BMI), tumor height, clinical T-stage, and clinical N-stage. The matching ratio was 1:1 and the caliper was 0.25 by default. All factors with significant differences in univariate analysis were included in the multivariate analysis and logistic regression was applied. The Kappa statistic was used to evaluate interobserver correlation between radiologists. Intraclass correlation coefficient (ICC) was used to evaluate consistency among the three radiologists.

Statistical analysis was carried out using SAS software (version 9.4; SAS Institute Inc., Cary, NC, USA). Statistical significance was set at p <0.05. All the work has been reported in line with the STROCSS criteria.(15)

## RESULTS

### Patients’ demographic and clinical characteristics at baseline

Patients with LARC who received CRT at our hospital between January 2000 and Aug 2021 were retrospectively analyzed. After exclusion, 1331 patients were included, 522 in the preoperative CRT group (Group 1) and 809 in the postoperative CRT group (Group 2). A flowchart of patient selection is shown in **Figure 2**. Of the 1331 patients, 578 patients underwent rectal MRI one year post-surgery (**Dataset 1**), including 271 and 307 patients in Group 1 and Group 2, respectively. **Dataset 1** was used to compare AFS and PFS (measured with MRI findings of the rectum one year post-surgery) between both groups. Of the 1331 patients, 883 were followed up for two years or longer (**Dataset 2**), including 415 and 468 patients in Group 1 and Group 2, respectively (**Figure 2**). **Dataset 2** was used to compare the incidence of ileostomy reversal and anorectal function at two years post-surgery between both groups (**Figure 2**).

**Figure 2.** The flowchart of patients’ selection.

### Comparison of clinicopathological parameters, AFS, and PFS between the two groups before matching in Dataset 1

The baseline clinicopathological parameters, AFS and PFS, of the two groups in Dataset 1 are presented in **Table 1**. Compared to Group 1, significantly greater tumor height, earlier clinical T-stage (c-T stage), higher pathological T-stage (p-T stage), higher pathological N-stage (p-N stage), and lower AFS and PFS were found in Group 2 (all P<0.05, **Table 1**).

### Comparison of clinicopathological parameters, AFS, and PFS between the two groups after matching in dataset 1

PSM with 130 pairs of patients was used to balance the influencing factors between the two groups, including age, gender, BMI, tumor height, c-T stage, and c-N stage. Although no significant difference was found for these parameters after matching, significantly higher p-T and p-N stages were found in Group 2 than in Group 1. This may have been related to the downstaging effect of preoperative CRT in Group 1 (**Table 1**). Significantly lower AFS and PFS were identified in Group 2 compared to Group 1 (**Table 1**).

### Univariate and multivariate analyses of possible risk factors of AFS in dataset 1

Univariate analysis showed that c-T stage, p-N stage, and timing of CRT were potential risk factors for AFS (all P<0.05, **Table 2**). In the following multivariate analysis, only the timing of CRT (P < 0.05) was an independent factor predicting AFS (odds ratio (OR): 0.098, 95% confidence interval (CI): 0.047-0.192).

### Univariate and multivariate analyses of possible risk factors of PFS in Dataset 1

Univariate analysis showed that c-T stage, p-T stage, p-N stage, and timing of CRT were potential risk factors for PFS (all P<0.05, **Table 2**). Multivariate analysis revealed that the timing of CRT was the only independent factor predicting PFS (OR: 0.165, 95%CI: 0.089-0.313).

### Comparison of clinicopathological parameters and the status of stoma closure before and after matching, between the two groups in Dataset 2

The clinicopathological parameters and the status of stoma closure in the two groups in Dataset 2 are summarized in **Table 3**. Compared to Group 1, Group 2 had significantly greater tumor height, lower c-T stage, lower p-T stage, lower p-N stage, lower incidence of temporary ileostomy (98.1% vs. 75.9%, P<0.01), and higher percentage of ileostomy reversal (65.4% vs. 88.2%, P<0.01, **Table 3**). After matching, a significantly lower and higher percentage of temporary ileostomy (98.5% vs. 87.7%, P<0.01) and ileostomy reversal (66.2% vs. 86.6%, P<0.01), respectively, were found in Group 2 than in Group 1 (**Table 3**, P<0.01). Importantly, anastomotic leakage (AL) incidence was greater in Group 1 (8.4%) than in Group 2 (4.9%), a statistically significant difference. After PSM, the incidence of AL remained greater in Group 1, indicating that neoadjuvant chemoradiotherapy potentially had adverse effects on anastomotic healing.

### Univariate and multivariate analyses of possible predictive factors of temporary ileostomy in Dataset 2

Univariate analysis showed that gender, age, tumor height, and timing of CRT correlated with the selection of temporary ileostomy (all P<0.05, **Table S2**). Multivariate analysis demonstrated that sex, age, tumor height, and timing of CRT (OR=0.06, 95%CI: 0.03-0.16, P<0.01) were independent predictors of temporary ileostomy (**Table S3**), and a significantly lower percentage of temporary ileostomy was found in Group 2 than in Group 1.

### Univariate and multivariate analyses of possible predictive factors of ileostomy reversal in Dataset 2

Patients with an ileostomy were recommended to under ileostomy reversal at 3-6 months after the first radical excision of rectal cancer in both groups. Univariate analysis showed that the c-T stage and timing of CRT were associated with ileostomy reversal (both P<0.05, **Table S4**). Multivariate analysis demonstrated that c-T stage and timing of CRT (OR=3.52, 95% CI: 2.71-5.40, P<0.01) were independent predictors of ileostomy reversal (**Table S5**). A significantly higher percentage of ileostomy reversal was noted in Group 2 than in Group 1.

### Comparison of Wexner score between the two groups in dataset 2

Of the 883 patients in Dataset 2, 121 had no temporary ileostomy whereas 762 did (579 of which underwent ileostomy reversal within two years) (**Table 3**). Therefore, anal sphincter function was assessed in 700 patients two years post-surgery and 532 completed the Wexner Fecal Incontinence Questionnaire. The clinicopathological features of the 532 patients are summarized in **Table S6**. There were no significant differences between two groups in pre-treatment Wexner scores. The mean Wexner score at two years after operation in Group 2 was significantly lower than that in Group 1 (5.3 vs. 2.3, P<0.01, **Table S6**). PSM was used to balance the following factors between both groups: age, sex, BMI, tumor height, c-T stage, and c-N stage. A total of 140 pairs of patients were successfully matched. After matching, the mean Wexner score in Group 2 was markedly lower than that in Group 1 (5.3 vs. 2.3, P<0.01, **Table S7**).

## DISCUSSION

We evaluated the influence of CRT timing on pelvic fibrosis and anorectal function in patients with LARC. Compared to the preoperative CRT group, the postoperative CRT group had higher rate of ileostomy reversal, and lower AFS, PFS, Wexner score, and percentage of temporary ileostomy. After matching, the results remained unchanged. Univariate and multivariate analyses demonstrated that the timing of CRT was an independent predictor of AFS, PFS, and rates of temporary ileostomy and ileostomy reversal. We also demonstrated that AFS and PFS positively correlated with Wexner scores. Taken together, our study suggested that postoperative CRT was associated with less pelvic fibrosis and better anorectal function than preoperative CRT in the treatment of patients with locally advanced rectal cancer.

In clinical practice, all patients with cT3-4 or lymph node metastasis (N+) rectal cancer were recommended to take preoperative CRT according to the NCCN guidelines. On the other hand, patients with early stage rectal cancer usually received operation directly, followed by postoperative CRT if they were staged as pT3-4 or N+ by postoperative pathological examination. For this reason, patients in Group 2 generally had earlier c-T and c-N stage than patients in Group1. In addition, the tumor in Group 1 will probably undergo down-staging after preoperative CRT, and this would result in the earlier p-T and p-N stage in Group 1. Although several studies have demonstrated that both preoperative and postoperative CRT may adversely affect anorectal function in patients with rectal cancer (16–21), the current consensus suggests neoadjuvant CRT to treat LARC. No study has investigated the differences in anorectal function between the two groups. To our knowledge, this is the first comparative study on pelvic fibrosis and anorectal function. Our study demonstrated that postoperative CRT might be superior to preoperative CRT in terms of severity of pelvic fibrosis and loss of anorectal function. It has been assumed that preoperative CRT preserves greater anorectal function than postoperative CRT; however, our results contradict this assumption, though with unknown underlying mechanism. Radiotherapy may potentially induce fibrosis, which is worsened by the following operative trauma, leading to worse anorectal function. Secondly, preoperative CRT also impairs leukocyte production and reduces collagen accumulation, leading to poor wound healing and an increased incidence of infection and anastomotic leakage (22). In our analysis, the preoperative CRT group had a higher incidence of temporary ileostomy. There is a growing body of data suggesting that, the duration from ileostomy to closure of ileostoma is associated with adverse anorectal function (23).

Our result might lead to the modification of clinical practice in the selection of CRT timing for some patients with LARC. Radiotherapy may induce necrosis, adhesion, fibrosis, and stenosis of the internal and external anal sphincter, puborectalis, and levator ani muscles (24, 25), thereby impairing anorectal function. At present, reliable parameters to evaluate fibrosis in the anastomosis, perirectal area, and pelvis have not been established. The radiological presentation of postoperative changes was typical. On T2WI scans, the anastomosis is reflected by a regular or irregular low-signal intensity layer localized in the rectal wall (hypointense similar to the muscularis propria layer), with covering by the normal mucosa. Simultaneously, postoperative anastomosis fibrosis tissue or scar proliferation is an inner thick layer or nodular focus of homogeneous intermediate-low signal intensity, which breaches or ruptures the hypointense layer, representing fibrotic scar tissue proliferation, with or without anastomotic stenosis. Perirectal fibrosis refers to peri-anastomotic, presacral and pelvic wall fibrous tissue hyperplasia and adhesion. The surrounding postoperative local-regional fibrosis tissue typically has a low-signal intensity on T2WI that included irregular linear or fibrous appearances. A second highlight of the current study is the use of AFS and PFS to quantify CRT-induced fibrosis in the anastomosis and perirectal area. Since both are related to the Wexner score, they can be used to assess the extent of pelvic fibrosis and anorectal function, and may also be applied in the mechanism and prevention research of CRT-induced fibrosis in the future.

Preoperative and postoperative CRT are both safe and promising treatments for patients with LARC, and the selection should be individualized according to the patient characteristics (26). Some patients with LARC could achieve clinical complete remission (cCR) or near-cCR after preoperative CRT. These patients have the possibility of receiving watch and wait strategy (W&W) or local resection to preserve the rectum, and their anorectal function would be better than patients who received postoperative CRT. But for those who still required radical excision after preoperative CRT, postoperative CRT is superior to preoperative CRT in terms of pelvic fibrosis and anorectal function, and the transition from preoperative CRT to postoperative CRT may lead to improvement in long-term anorectal function. However, preoperative CRT still surpasses postoperative CRT in terms of rates of local recurrence, percentage of patients requiring radical excision, and sphincter preservation. Therefore, it is highly important to screen patients who may benefit from preoperative CRT, including patients at high risk of local recurrence, extramural vascular invasion (EMVI), mesorectal fascia (MRF) involvement, low-lying rectal cancer, and large and non-resectable tumors (27). As the accuracy of preoperative MRI for determining TNM stage is only about 60-80% (28), preoperative CRT may lead to under-treatment in some patients and over-treatment in others. The selection of postoperative CRT is based on intraoperative exploration and postoperative pathological examination, which is noticeably more accurate (3). In the United States, only 50% of patients with LARC have received preoperative CRT (29). The transition from preoperative CRT to postoperative CRT may improve treatment accuracy and long-term anorectal function for selected patients. Postoperative CRT can be selectively used in patients with adverse histopathological features (30). It may also lead to avoidance of unnecessary radiotherapy for patients with a false preoperative advanced stage, resulting in better anorectal function.

This study’s limitations are as follows: Firstly, it was a retrospective single-center study, thus, selection and recall biases were inevitable; Secondly, there were significant differences in tumor height and clinical T-stage between both groups, which may interfere with their comparisons. In this study, multivariate analysis and PSM were used to minimize the interference of the unbalanced baseline. Thirdly, both surgical approaches and radiotherapy equipment and methods may have advanced since the time of this study, limiting its application to real-world settings.

In conclusion, AFS and PFS were introduced to evaluate CRT-induced fibrosis. Our results demonstrated that postoperative CRT surpassed preoperative CRT in terms of AFS, PFS, percentage of temporary ileostomy and ileostomy reversal, and long-term anorectal function. Our findings may assist clinicians in optimizing the sequence of radiotherapy for treating patients with LARC. Our results suggest that preoperative CRT might not be the optimal treatment for some patients with LARC in terms of anorectal function, especially in patients with a low risk of local recurrence. Instead, the selection of postoperative CRT based on postoperative pathological examination would be more accurate and have better anorectal function. However, as shown in Table 1 and Table 3, the tumors might shrink significantly after preoperative CRT, which allowed some patients with unresectable tumors to undergo surgery. This should be taken into consideration in the decision making between preoperative or postoperative CRT. In addition, the underlying reason for worse fibrosis and anorectal function in the preoperative CRT remains elusive, and further prospective RCTs with a larger sample size are required to validate these findings.

## Data Availability

All data produced in the present study are available upon reasonable request to the authors

## Reference

1. Sauer R, Becker H, Hohenberger W, Rodel C, Wittekind C, Fietkau R, et al. Preoperative versus postoperative chemoradiotherapy for rectal cancer. N Engl J Med. 2004;351(17):1731–40.

2. Krook JE, Moertel CG, Gunderson LL, Wieand HS, Collins RT, Beart RW, et al. Effective surgical adjuvant therapy for high-risk rectal carcinoma. N Engl J Med. 1991;324(11):709–15.

3. Kim JH. Controversial issues in radiotherapy for rectal cancer: a systematic review. Radiat Oncol J. 2017;35(4):295–305.

4. Song JH, Jeong JU, Lee JH, Kim SH, Cho HM, Um JW, et al. Preoperative chemoradiotherapy versus postoperative chemoradiotherapy for stage II-III resectable rectal cancer: a meta-analysis of randomized controlled trials. Radiat Oncol J. 2017;35(3):198–207.

5. Sauer R, Liersch T, Merkel S, Fietkau R, Hohenberger W, Hess C, et al. Preoperative versus postoperative chemoradiotherapy for locally advanced rectal cancer: results of the German CAO/ARO/AIO-94 randomized phase III trial after a median follow-up of 11 years. J Clin Oncol. 2012;30(16):1926–33.

6. NCCN clincal practice guidelines in onology, version 1.2020-December 19, 2019. rectal cancer, available at: https://www.nccn.org/professionals/physician_gls/pdf/rectal.pdf.

7. Lim YJ, Kim Y, Kong M. Comparative survival analysis of preoperative and postoperative radiotherapy in stage II-III rectal cancer on the basis of long-term population data. Sci Rep. 2018;8(1):17153.

8. Marinello FG, Frasson M, Baguena G, Flor-Lorente B, Cervantes A, Rosello S, et al. Selective approach for upper rectal cancer treatment: total mesorectal excision and preoperative chemoradiation are seldom necessary. Dis Colon Rectum. 2015;58(6):556–65.

9. Zhu X, Lou Z, Gong H, Meng R, Hao L, Zhang W. Influence of neoadjuvant chemoradiotherapy on the anal sphincter: ultrastructural damage may be critical. Int J Colorectal Dis. 2016;31(8):1427–30.

10. Wang B, Wei J, Meng L, Wang H, Qu C, Chen X, et al. Advances in pathogenic mechanisms and management of radiation-induced fibrosis. Biomed Pharmacother. 2020;121:109560.

11. Matzel K, Bittorf B, Günther K, Stadelmaier U, Hohenberger W. Rectal resection with low anastomosis: functional outcome. Colorectal Dis. 2003;5(5):458–64.

12. Lundby L, Krogh K, Jensen V, Gandrup P, Qvist N, Overgaard J, et al. Long-term anorectal dysfunction after postoperative radiotherapy for rectal cancer. Diseases of the colon and rectum. 2005;48(7):1343–9; discussion 9-52; author reply 52.

13. Yuan Y, Yu Y, Sun Y, Li S, Lu H, Ma X, et al. Investigating anorectal function using postoperative MRI-based fibrosis score in patients with locally advanced rectal cancer receiving neoadjuvant chemoradiotherapy: a two-center study. Annals of medicine. 2023;55(2):2268112.

14. Essangri H, Majbar M, Benkabbou A, Amrani L, Mohsine R, Souadka A. Transcultural adaptation and validation of the Moroccan Arabic dialect version of the Wexner incontinence score in patients with low anterior resection syndrome after rectal surgery. Surgery. 2021;170(1):47–52.

15. Mathew G and Agha R for the STROCSS Group. STROCSS 2021: Strengthening the reporting of cohort, cross-sectional and case-control studies in Surgery. International journal of surgery 2021;96:106165.

16. Lim J, Tjandra J, Hiscock R, Chao M, Gibbs P. Preoperative chemoradiation for rectal cancer causes prolonged pudendal nerve terminal motor latency. Dis Colon Rectum 2006;49(1):12–9.

17. Gervaz P, Rotholtz N, Pisano M, Kaplan E, Secic M, Coucke P, et al. Quantitative short-term study of anal sphincter function after chemoradiation for rectal cancer. Arch Surg. 2001;136(2):192–6.

18. Ammann K, Kirchmayr W, Klaus A, Mühlmann G, Kafka R, Oberwalder M, et al. Impact of neoadjuvant chemoradiation on anal sphincter function in patients with carcinoma of the midrectum and low rectum. Arch Surg. 2003;138(3):257–61.

19. Canda A, Terzi C, Gorken I, Oztop I, Sokmen S, Fuzun M. Effects of preoperative chemoradiotherapy on anal sphincter functions and quality of life in rectal cancer patients. Int J Colorectal Dis 2010;25(2):197–204.

20. De Nardi P, Testoni S, Corsetti M, Andreoletti H, Giollo P, Passaretti S, et al. Manometric evaluation of anorectal function in patients treated with neoadjuvant chemoradiotherapy and total mesorectal excision for rectal cancer. Dig Liver Dis. 2017;49(1):91–7.

21. Ihnát P, Slívová I, Tulinsky L, Ihnát Rudinská L, Máca J, Penka I. Anorectal dysfunction after laparoscopic low anterior rectal resection for rectal cancer with and without radiotherapy (manometry study). J Surg Oncol 2018;117(4):710–6.

22. Johnson LB, Jorgensen LN, Adawi D, Blomqvist P, Asklof GB, Gottrup F, et al. The effect of preoperative radiotherapy on systemic collagen deposition and postoperative infective complications in rectal cancer patients. Dis Colon Rectum. 2005;48(8):1573–80.

23. Gadan S, Floodeen H, Lindgren R, Matthiessen P. Does a Defunctioning Stoma Impair Anorectal Function After Low Anterior Resection of the Rectum for Cancer? A 12-Year Follow-up of a Randomized Multicenter Trial. Dis Colon Rectum 2017;60(8):800–6.

24. Lakhoo J, Khatri G, Elsayed R, Chernyak V, Olpin J, Steiner A, et al. MRI of the Male Pelvic Floor. Radiographics. 2019;39(7):2003–22.

25. Khatri G, de Leon A, Lockhart M. MR Imaging of the Pelvic Floor. Magn Reson Imaging Clin N Am. 2017;25(3):457–80.

26. Lee BC, Park IJ, Kim CW, Lim SB, Yu CS, Kim JC. Matched case-control analysis comparing oncologic outcomes between preoperative and postoperative chemoradiotherapy for rectal cancer. Ann Surg Treat Res. 2017;92(4):200–7.

27. Glimelius BL. The role of preoperative and postoperative radiotherapy in rectal cancer. Clin Colorectal Cancer. 2002;2(2):82–92.

28. Kim NK, Kim MJ, Park JK, Park SI, Min JS. Preoperative staging of rectal cancer with MRI: accuracy and clinical usefulness. Ann Surg Oncol. 2000;7(10):732–7.

29. Sineshaw HM, Jemal A, Thomas CR, Jr., Mitin T. Changes in treatment patterns for patients with locally advanced rectal cancer in the United States over the past decade: An analysis from the National Cancer Data Base. Cancer. 2016;122(13):1996–2003.

30. Glynne-Jones R, Wyrwicz L, Tiret E, Brown G, Rodel C, Cervantes A, et al. Rectal cancer: ESMO clinical practice guidelines for diagnosis, treatment and follow-up. Ann Oncol. 2017;28(suppl_4):iv22–iv40.

