## Supplementary material for "Postoperative Chemoradiotherapy causes Less Fibrosis and Better Anorectal Function compared with preoperative: A retrospective study in Rectal Cancer": table1-table3 (from another version of this preprint: version 1, 2024-11-29)

**Tables**

**Table 1**. Clinicopathological parameters, AFS, and PFS of both groups before and after matching in Dataset 1

| Clinicopathological parameters | | Before matching | | | After matching | | |
| --- | --- | --- | --- | --- | --- | --- | --- |
|  |  | Group 1  (n=133) | Group 2  (n=207) | *P*-value | Group 1  (n=120) | Group 2  (n=120) | *P*-value |
| Gender |  |  |  | 0.07 |  |  | 0.62 |
|  | Male | 101(75.9) | 138(66.7) |  | 72(74.2) | 75(77.3) |  |
|  | Female | 32(24.1) | 69(33.3) |  | 25(25.8) | 22(22.7) |  |
| Age (years) | x̅ ±s | 56.9±11.4 | 58.0±10.6 | 0.36 | 57.3±11.1 | 58.0±11.3 | 0.64 |
| BMI | x̅ ±s | 23.2±3.2 | 23.3±2.9 | 0.70 | 23.3±3.0 | 23.3±3.2 | 0.99 |
| Tumor height |  |  |  | <0.01 |  |  | 0.72 |
|  | ≤5cm | 113(85.0) | 134(64.7) |  | 78(80.4) | 76(78.4) |  |
|  | >5cm | 20(15.0) | 73(35.3) |  | 19(19.6) | 21(21.6) |  |
| c-T stage |  |  |  | 0.01 |  |  | 1.00 |
|  | T1-2 | 18(15.0) | 59(28.5) |  | 18(18.6) | 18(18.6) |  |
|  | T3-4 | 102(85.0) | 148(71.5) |  | 79(81.4) | 79(81.4) |  |
| c-N stage |  |  |  | 0.08 |  |  | 0.75 |
|  | N0-1 | 89(74.2) | 134(64.7) |  | 67(69.1) | 69(71.1) |  |
|  | N2 | 31(25.8) | 73(35.3) |  | 30(30.9) | 28(28.9) |  |
| p-T stage |  |  |  | <0.01 |  |  | <0.01 |
|  | T0-2 | 71(53.8) | 44(21.9) |  | 53(55.2) | 18(18.9) |  |
|  | T3-4 | 61(46.2) | 157(78.1) |  | 43(44.8) | 77(81.1) |  |
| p-N stage |  |  |  | <0.01 |  |  | <0.01 |
|  | N0-1 | 121(91.7) | 120(59.7) |  | 87(90.6) | 61(64.2) |  |
|  | N2 | 11(8.3) | 81(40.3) |  | 9(9.4) | 34(35.8) |  |
| AFS |  |  |  | <0.01 |  |  | <0.01 |
|  | 0 | 1(0.8) | 51(24.6) |  | 0(0.0) | 22(22.7) |  |
|  | 1 | 67(50.4) | 137(66.2) |  | 48(49.5) | 63(64.9) |  |
|  | 2 | 65(48.9) | 19(9.2) |  | 49(50.5) | 12(12.4) |  |
| PFS |  |  |  | <0.01 |  |  | <0.01 |
|  | 0 | 3(2.3) | 26(12.6) |  | 3(3.1) | 14(14.4) |  |
|  | 1 | 72(54.1) | 161(77.8) |  | 53(54.6) | 71(73.2) |  |
|  | 2 | 58(43.6) | 20(9.7) |  | 41(42.3) | 12(12.4) |  |

BMI: body mass index; AFS, anastomotic fibrosis score; PFS, perirectal fibrosis score**;** Group 1, preoperative CRT group; Group 2, postoperative CRT group;

**Table 2.** Univariate analysis of possible risk factors of AFS and PFS in Dataset 1

| Clinicopathological parameters | | Overall  (n=340) | AFS | | | | PFS | | | |
| --- | --- | --- | --- | --- | --- | --- | --- | --- | --- | --- |
|  |  |  | 0 (n=52) | 1 (n=204) | 2 (n=84) | *P*-value | 0 (n=29) | 1 (n=233) | 2 (n=78) | *P*-value |
| Gender |  |  |  |  |  | 0.29 |  |  |  | 0.17 |
|  | Male | 239 (70.3) | 32 (61.5) | 145 (71.1) | 62 (73.8) |  | 18 (62.1) | 160 (68.7) | 61 (78.2) |  |
|  | Female | 101 (29.7) | 20 (38.5) | 59 (28.9) | 22 (26.2) |  | 11 (37.9) | 73 (31.3) | 17 (21.8) |  |
| Age (years) | x̅ ±s | 57.6±10.9 | 55.6±10.3 | 58.4±10.9 | 56.6±11.4 | 0.21 | 55.2±10.5 | 58.4±10.8 | 56.1±11.3 | 0.14 |
| BMI | x̅ ±s | 23.3±3.0 | 22.9±3.0 | 23.3±3.2 | 23.4±2.7 | 0.57 | 23.2±3.1 | 23.3±3.0 | 23.3±3.1 | 0.98 |
| Tumor height |  |  |  |  |  | 0.07 |  |  |  | 0.09 |
|  | ≤5cm | 247 (72.6) | 41 (78.8) | 139 (68.1) | 67 (79.8) |  | 19 (65.5) | 164 (70.4) | 64 (82.1) |  |
|  | >5cm | 93 (27.4) | 11 (21.2) | 65 (31.9) | 17 (20.2) |  | 10 (34.5) | 69 (29.6) | 14 (17.9) |  |
| c-T stage |  |  |  |  |  | 0.04 |  |  |  | <0.01 |
|  | T1-2 | 77 (23.5) | 19 (37.3) | 44 (22.0) | 14 (18.4) |  | 6 (20.7) | 64 (28.2) | 7 (9.9) |  |
|  | T3-4 | 250 (76.5) | 32 (62.7) | 156 (78.0) | 62 (81.6) |  | 23 (79.3) | 163 (71.8) | 64 (90.1) |  |
| c-N stage |  |  |  |  |  | 0.22 |  |  |  | 0.26 |
|  | N0-1 | 223 (68.2) | 33 (64.7) | 132 (66.0) | 58 (76.3) |  | 16 (55.2) | 156 (68.7) | 51 (71.8) |  |
|  | N2 | 104 (31.8) | 18 (35.3) | 68 (34.0) | 18 (23.7) |  | 13 (44.8) | 71 (31.3) | 20 (28.2) |  |
| p-T stage |  |  |  |  |  | 0.19 |  |  |  | 0.02 |
|  | T0-2 | 115 (34.5) | 18 (36.0) | 62 (31.0) | 35 (42.2) |  | 5 (17.2) | 76 (33.5) | 34 (44.2) |  |
|  | T3-4 | 218 (65.5) | 32 (64.0) | 138 (69.0) | 48 (57.8) |  | 24 (82.8) | 151 (66.5) | 43 (55.8) |  |
| p-N stage |  |  |  |  |  | <0.01 |  |  |  | 0.02 |
|  | N0-1 | 241 (72.4) | 32 (64.0) | 136 (68.0) | 73 (88.0) |  | 16 (55.2) | 162 (71.4) | 63 (81.8) |  |
|  | N2 | 92 (27.6) | 18 (36.0) | 64 (32.0) | 10 (12.0) |  | 13 (44.8) | 65 (28.6) | 14 (18.2) |  |
| Timing of CRT | |  |  |  |  | <0.01 |  |  |  | <0.01 |
|  | Group 1 | 133 (39.1) | 1 (1.9) | 67 (32.8) | 65 (77.4) |  | 3 (10.3) | 72 (30.9) | 58 (74.4) |  |
|  | Group 2 | 207 (60.9) | 51 (98.1) | 137 (67.2) | 19 (22.6) |  | 26 (89.7) | 161 (69.1) | 20 (25.6) |  |

BMI: body mass index; CRT: chemoradiation therapy; AFS, anastomotic fibrosis score; PFS, perirectal fibrosis score; Group 1, preoperative CRT group; Group 2, postoperative CRT group;

**Table 3**. Clinicopathological parameters, stoma closure status, and Wexner score of both groups before and after matching in Dataset 2.

| Clinicopathological parameters | | Before matching | | | After matching | | |
| --- | --- | --- | --- | --- | --- | --- | --- |
|  |  | Group 1  (n=247) | Group 2  (n=315) | *P*-value | Group 1  (n=182) | Group2  (n=182) | P value |
| Gender |  |  |  | 0.08 |  |  | 0.74 |
|  | Male | 171(69.2) | 196(62.2) |  | 123(67.6) | 120(65.9) |  |
|  | Female | 76(30.8) | 119(37.8) |  | 59(32.4) | 62(34.1) |  |
| Age (years) | x̅ ±s | 56.2±11.5 | 56.9±11.5 | 0.43 | 56.2±11.2 | 56.6±11.8 | 0.72 |
| BMI | x̅ ±s | 23.3±3.3 | 23.5±3.1 | 0.47 | 23.4±3.3 | 23.5±3.2 | 0.86 |
| Tumor height | |  |  | <0.01 |  |  | 0.90 |
|  | ≤5cm | 197(79.8) | 183(58.1) |  | 140(76.9) | 139(76.4) |  |
|  | >5cm | 50(20.2) | 132(41.9) |  | 42(23.1) | 43(23.6) |  |
| c-T stage |  |  |  | 0.01 |  |  | 1.00 |
|  | T1-2 | 35(16.7) | 73(26.4) |  | 34(18.7) | 34(18.7) |  |
|  | T3-4 | 174(83.3) | 204(73.6) |  | 148(81.3) | 148(81.3) |  |
| c-N stage |  |  |  | 0.58 |  |  | 0.74 |
|  | N0-1 | 140(67.0) | 192(69.3) |  | 120(65.9) | 117(64.3) |  |
|  | N2 | 69(33.0) | 85(30.7) |  | 62(34.1) | 65(35.7) |  |
| p-T stage |  |  |  | <0.01 |  |  | <0.01 |
|  | T0-2 | 125(52.3) | 58(21.3) |  | 93(51.7) | 34(18.9) |  |
|  | T3-4 | 114(47.7) | 214(78.7) |  | 87(48.3) | 146(81.1) |  |
| p-N stage |  |  |  | <0.01 |  |  | <0.01 |
|  | N0-1 | 223(93.3) | 176(64.7) |  | 167(92.8) | 110(61.1) |  |
|  | N2 | 16(6.7) | 96(35.3) |  | 13(7.2) | 70(38.9) |  |
| Temporary ileostomy | |  |  | <0.01 |  |  | <0.01 |
|  | No | 5(2.0) | 76(24.1) |  | 2(1.1) | 22(12.1) |  |
|  | Yes | 242(98.0) | 239(75.9) |  | 180(98.9) | 160(87.9) |  |
| Stoma closure | |  |  | <0.01 |  |  | <0.01 |
|  | Unclosed | 84(34.7) | 28(11.7) |  | 61(33.9) | 21(13.1) |  |
|  | Closed | 158(65.3) | 211(88.3) |  | 119(66.1) | 139(86.9) |  |
| Anastomotic Leakage | |  |  |  |  |  |  |
|  | No | 226(91.5) | 299(95.0) | <0.05 | 166(91.2) | 173(95.1) | <0.05 |
|  | Yes | 21(8.5) | 16(5.0) |  | 16(8.8) | 9(4.9) |  |

BMI: body mass index; Group 1: preoperative CRT group; Group 2: postoperative CRT group.
