## supplemental figures and tables for "Postoperative Chemoradiotherapy causes Less Fibrosis and Better Anorectal Function compared with preoperative: A retrospective study in Rectal Cancer" (from another version of this preprint: version 1, 2024-11-29)

**Supplementary figures and tables**

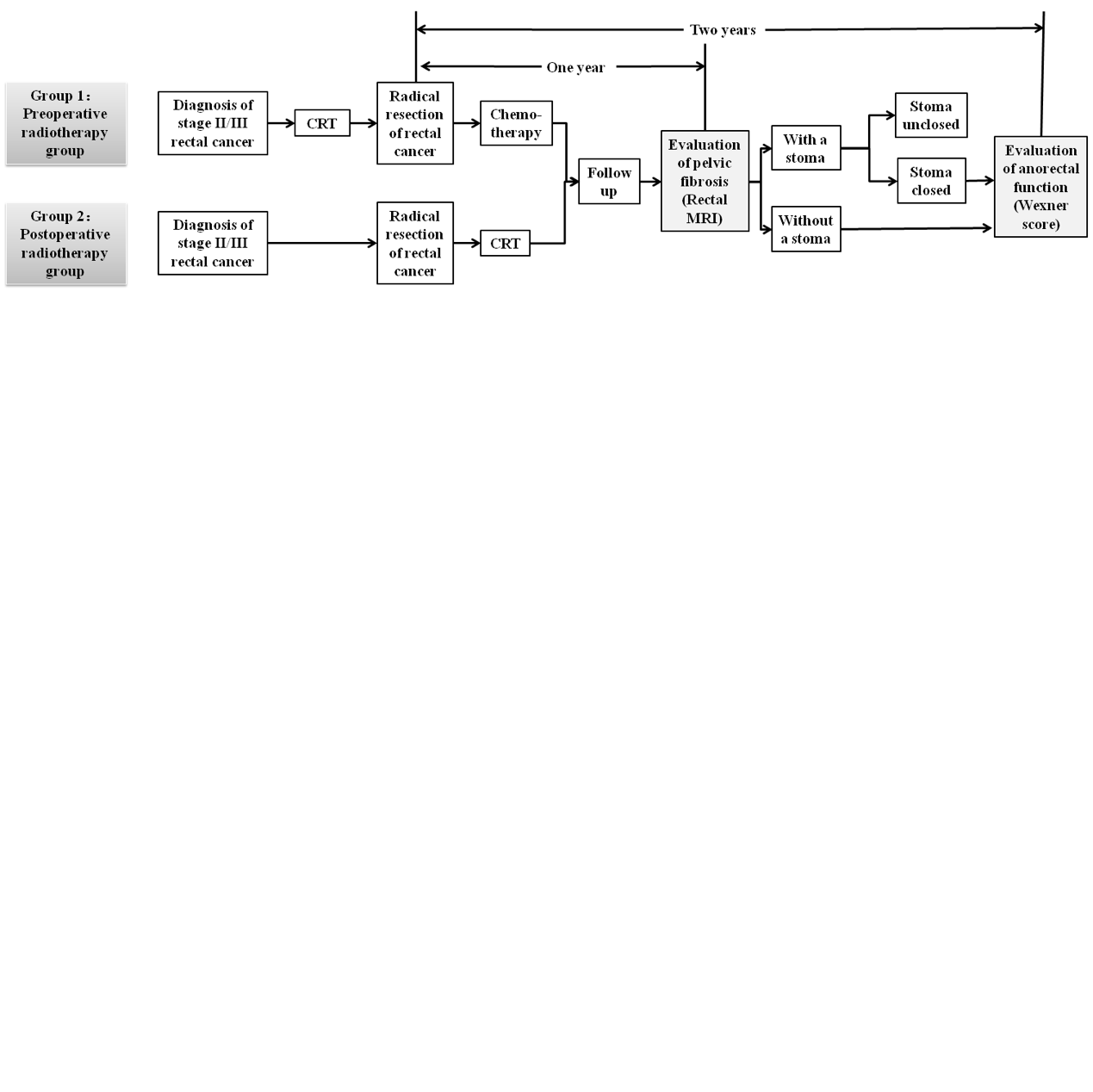

**Figure S1.** Treatment process and evaluation of pelvic fibrosis (based on rectal MRI) and anorectal function (Wexner score) in the two groups. CRT: chemoradiotherapy.

**Supplementary Table 1.** Interobserver agreement for MR fibrosis scores

| Cohort | Score | Radiologist 1 - Radiologist 2  *Kappa* value (95% CI) | Radiologist 1 - Senior Radiologist  *Kappa* value (95% CI) | Radiologist 2 - Senior Radiologist  *Kappa* value (95% CI) | All Radiologists  ICC (95% CI) |
| --- | --- | --- | --- | --- | --- |
| Group 1 | AFS | 0.863 (0.715 - 0.950) | 0.918 (0.873 - 0.954) | 0.863 (0.826 - 0.973) | 0.873 (0.836 - 0.947) |
|  | PFS | 0.663 (0.574 - 0.847) | 0.827 (0.745 - 0.974) | 0.847 (0.757 - 0.945) | 0.773 (0.747 - 0.854) |
| Group 2 | AFS | 0.836 (0.773 - 0.956) | 0.916 (0.873 - 0.971) | 0.904 (0.873 - 0.979) | 0.873 (0.832 - 0.947) |
|  | PFS | 0.736 (0.663 - 0.873) | 0.846 (0.757 - 0.916) | 0.890 (0.834 - 0.942) | 0.803 (0.743 - 0.873) |

ICC: intraclass correlation coefficient; AFS: anastomotic fibrosis score; PFS: perirectal fibrosis score.

**Supplementary Table 2.** Univariate analysis of predictive factors of preventive ileostomy in dataset 2.

| Clinicopathological parameters | | Without a stoma  (n=121) | With a stoma  (n=762) | P-value |
| --- | --- | --- | --- | --- |
| Gender | |  |  | <0.01 |
|  | Male | 64(52.9) | 514(67.5) |  |
|  | Female | 57(47.1) | 248(32.5) |  |
| Age (years) | x̅ ±s | 52.4±11.8 | 57.6±10.3 | 0.03 |
| BMI | x̅ ±s | 23.2±3.7 | 23.7±3.1 | 0.15 |
| Tumor height |  |  |  | <0.01 |
|  | ≤5cm | 31(25.6) | 568(74.5) |  |
|  | >5cm | 90(74.4) | 194(25.5) |  |
| c-T stage |  |  |  | 0.46 |
|  | T1-2 | 33(27.3) | 165(21.7) |  |
|  | T3-4 | 88(72.7) | 597(78.3) |  |
| c-N stage |  |  |  | 0.61 |
|  | N0-1 | 88(72.7) | 516(67.7) |  |
|  | N2 | 33(27.3) | 246(32.3) |  |
| Timing of CRT | |  |  | <0.01 |
|  | Group1 | 8(6.6) | 383(50.3) |  |
|  | Group2 | 113(93.4) | 379(49.7) |  |

BMI: body mass index; CRT: chemoradiation therapy; Group 1: preoperative CRT group, Group 2: postoperative CRT group.

**Supplementary Table 3.** Multivariate analysis of predictive factors of preventive ileostomy in dataset 2.

|  | OR | 95%CI | *P value* |
| --- | --- | --- | --- |
| Gender (Female vs. Male) | 0.41 | 0.22-0.79 | 0.01 |
| Age (＞65y vs. ≤65y) | 1.06 | 1.04-1.11 | ＜0.01 |
| Tumor height (＞5cm vs. ≤5cm) | 0.13 | 0.07-0.24 | ＜0.01 |
| Timing of CRT (Group 2 vs. Group 1) | 0.06 | 0.03-0.16 | ＜0.01 |

CRT: chemoradiation therapy

**Supplementary Table 4.** Univariate analysis of predictive factors of ileostoma reversal in dataset 2.

| Clinicopathological parameters | | Stoma unclosed  (n=183) | Stoma closed  (n=579) | *P value* |
| --- | --- | --- | --- | --- |
| Gender |  |  |  | 0.44 |
|  | Male | 118(64.5) | 395(68.2) |  |
|  | Female | 65(35.5) | 184(31.8) |  |
| Age (years) | x̅ ±s | 57.4±11.7 | 57.4±10.8 | 0.93 |
| BMI | x̅ ±s | 23.5±3.0 | 23.4±3.1 | 0.89 |
| Tumor height |  |  |  | 0.08 |
|  | ≤5cm | 149(81.4) | 420(72.5) |  |
|  | >5cm | 34(18.6) | 159(27.5) |  |
| c-T stage |  |  |  | <0.01 |
|  | T1-2 | 23(12.6) | 141(24.3) |  |
|  | T3-4 | 160(87.4) | 438(75.7) |  |
| c-N stage |  |  |  | 0.12 |
|  | N0-1 | 111(60.7) | 404(69.8) |  |
|  | N2 | 72(39.3) | 175(30.2) |  |
| p-T stage |  |  |  | 0.77 |
|  | T0-2 | 72(39.3) | 218(37.7) |  |
|  | T3-4 | 111(60.7) | 361(62.3) |  |
| p-N stage |  |  |  | 0.34 |
|  | N0-1 | 150(82.0) | 449(77.5) |  |
|  | N2 | 33(18.0) | 130(22.5) |  |
| Timing of CRT |  |  |  | <0.01 |
|  | Group1 | 137(74.9) | 248(42.8) |  |
|  | Group2 | 46(25.1) | 331(57.2) |  |

BMI: body mass index; CRT: chemoradiation therapy; Group 1: preoperative CRT group, Group 2: postoperative CRT group.

**Supplementary Table 5.** Multivariate analysis of predictive factors of ileostomy reversal in dataset 2.

|  | OR | 95%CI | *P* |
| --- | --- | --- | --- |
| c-T stage (T3-4 vs. T1-2) | 0.51 | 0.24-0.98 | 0.04 |
| Timing of CRT (Group 2 vs. Group 1) | 3.52 | 2.71-5.40 | ＜0.01 |

CRT: chemoradiation therapy; Group 1: preoperative CRT group, Group 2: postoperative CRT group.

**Supplementary Table 6.** Clinicopathological features and anal anorectal function of the 532 patients who had functional anus and had completed the Wexner Fecal Incontinence Questionnaire in dataset 2.

| Clinicopathological parameters | | Group 1  (n=188) | Group 2  (n=344) | P-value |
| --- | --- | --- | --- | --- |
| Gender |  |  |  | 0.08 |
|  | Male | 133(70.7) | 213(61.9) |  |
|  | Female | 55(29.3) | 131(38.1) |  |
| Age (years) | x̅±s | 55.9±10.8 | 56.7±11.4 | 0.45 |
| BMI | x̅±s | 23.4±3.1 | 23.3±2.8 | 0.93 |
| Tumor height |  |  |  | <0.01 |
|  | ≤5cm | 144(76.6) | 188(54.7) |  |
|  | >5cm | 44(23.4) | 156(45.3) |  |
| c-T stage |  |  |  | 0.20 |
|  | T1-2 | 38(20.2) | 93(27.0) |  |
|  | T3-4 | 150(79.8) | 251(73.0) |  |
| c-N stage |  |  |  | 0.82 |
|  | N0-1 | 136(72.3) | 241(70.1) |  |
|  | N2 | 52(27.7) | 103(29.9) |  |
| p-T stage |  |  |  | <0.01 |
|  | T0-2 | 104(55.3) | 76(22.1) |  |
|  | T3-4 | 76(44.7) | 268(77.9) |  |
| p-N stage |  |  |  | <0.01 |
|  | N0-1 | 177(94.1) | 228(66.3) |  |
|  | N2 | 11(5.9) | 116(33.7) |  |
| Wexner score* | After surgery | 5.3±3.4 | 2.3±2.1 | <0.01 |
|  | Pre-treatment | 1.2±0.1 | 1.3±0.1 | 0.95 |

* Wexner scores were recorded at pre-treatment and 2 years after surgery;

BMI: body mass index; Group 1: preoperative CRT group; Group 2: postoperative CRT group.

**Supplementary Table 7.** Clinicopathological parameters of the 125 pairs of patients with functional anus and Wexner scores at 2 years after surgery after matching in dataset 2

| Clinicopathological parameters | | Group1  (n=140) | Group2  (n=140) | *P-value* |
| --- | --- | --- | --- | --- |
| Gender |  |  |  | 0.77 |
|  | Male | 102(72.9) | 99 (70.7) |  |
|  | Female | 38(27.1) | 41 (29.3) |  |
| Age (years) | x̅ ±s | 56.7±10.5 | 56.6±10.3 | 0.99 |
| BMI | x̅ ±s | 23.1±3.7 | 23.3±2.5 | 0.87 |
| Tumor height |  |  |  | 0.73 |
|  | ≤5cm | 108(77.1) | 109(77.9) |  |
|  | >5cm | 32(22.9) | 31(22.1) |  |
| c-T stage |  |  |  | 0.14 |
|  | T1-2 | 28(17.9) | 32(22.9) |  |
|  | T3-4 | 112(87.1) | 108(77.1) |  |
| c-N stage |  |  |  | 0.86 |
|  | N0-1 | 100(71.4) | 99(70.7) |  |
|  | N2 | 40(28.6) | 41(29.3) |  |
| p-T stage |  |  |  | <0.01 |
|  | T0-2 | 76(54.3) | 29(20.7) |  |
|  | T3-4 | 64(45.7) | 111(79.3) |  |
| p-N stage |  |  |  | <0.01 |
|  | N0-1 | 132(94.3) | 93(66.4) |  |
|  | N2 | 8(5.7) | 47(33.6) |  |
| Wexner score | x̅ ±s | 5.3±2.5 | 2.3±1.7 | <0.01 |

BMI: body mass index; Group 1: preoperative CRT group, Group 2: postoperative CRT group.
